# SHEAR Saliva Collection Device Augments Sample Properties for Improved Analytical Performance

**DOI:** 10.1101/2022.07.07.22277204

**Authors:** Shang Wei Song, Rashi Gupta, Jothilingam Niharika, Xinlei Qian, Yue Gu, V Vien Lee, Yoann Sapanel, David Michael Allen, John Eu Li Wong, Paul A MacAry, Dean Ho, Agata Blasiak

## Abstract

Despite human saliva representing a convenient and non-invasive clinical substrate for disease diagnosis and biomonitoring, its widespread utilization has been hampered by technical challenges. The non-Newtonian, heterogenous and highly viscous nature of clinical saliva samples complicate the development of automated fluid handling processes that are vital for accurate diagnoses. Furthermore, conventional saliva processing methods are either resource and/or time intensive precluding certain testing capabilities in low- and middle-income countries, with these challenges aggravated during a pandemic outbreak. The conventional approaches can also potentially alter analyte structure, reducing application opportunities in Point-of-Care diagnostics. To overcome these challenges, we introduce the SHEAR saliva collection device that preprocesses saliva for enhanced interfacing with downstream assays. We demonstrate the device’s impact on reducing saliva’s viscosity, improving sample uniformity and, increasing diagnostic performance of COVID-19 Rapid Antigen Tests. Importantly, in addition to reporting technical advances and to address downstream implementation factors, we conducted a formal user experience study, which resulted in generally positive comments. Effective implementation of this device could be of support to realize the potential of saliva, particularly in large-scale and/or resource-limited settings for global and community health diagnostics.

## Introduction

Saliva is receiving increasing attention as an analytical sample for biomonitoring particularly in the context of the COVID-19 pandemic. In order to improve diagnostic and screening throughput among the other factors, saliva testing has been extensively evaluated. Saliva-based diagnostic testing for the detection of SARS-CoV-2 virus has been progressively adopted, including in the United States, Australia, and Singapore (*1-3*). The simple and non-invasive collection methods of saliva enable self-testing as sampling can be conducted safely without the supervision of trained personnel, reducing the risk of transmission of the pathogen to healthcare workers. Self-sample collection and testing allows for a decrease in manpower requirements, an increase in testing capacity and the potential for earlier identification/isolation of pre-symptomatic individuals. These are important considerations for all regions where initial outbreaks may strain testing resources and/or low- and middle-income countries (LMICs). SARS-CoV-2 RNA in saliva has been shown to remain stable at room temperature for more than a week (*4, 5*), eliminating the need for cold chain management. Recent studies reported enhanced detection of Omicron variant in saliva samples possibly due to higher viral shedding (*6*), suggesting the need to review the current diagnostic testing standard-of-care (SOC). Beyond SARS-CoV-2 testing, saliva has shown promise for other diagnostic applications due to the plethora of analytes such as hormones, enzymes and antibodies that it contains (*7*).

Beyond the analytes of current diagnostic interest, saliva contains a high concentration of glycoproteins, especially mucin (*7, 8*). Glycoproteins provide protection to buccal epithelium from chemicals, microbes and wear-and-tear but make saliva a difficult analytical matrix due to formation of pockets of high viscosity (*9*). Viscous matrix is troublesome as it complicates accurate sample processing (e.g. pipetting of accurate volumes) and automation (*10-14*). Additionally, salivary mucin networks form complexes with some of the proteins (*15*), potentially reducing their availability for detection. Furthermore, the pockets of viscosity result in non-uniform physical properties and non-uniform analyte distribution within the sample contributing to analytical variability. The current methods to homogenize biofluids include mechanical and chemical approaches, each with their own benefits and disadvantages. For example, a traditional mechanical method involves exposing the sample to cycles of freeze-thawing. Whilst this approach is cost-effective and does not require extensive laboratory equipment or reagents, it is time consuming and multiple cycles of freeze-thawing can alter the concentration of analyte (*16*). A new mechanical homogenization method was proposed using magnetic rods (*14*). While faster, it requires a centrifuge and laboratory equipment, and is not suitable for small volumes. Chemical methods are much more popular, among them mucinase (*17*) and dithiothreitol (DTT) (*18, 19*) have shown to degrade mucin, lowering sample’s viscosity. While relatively fast (<1h incubation) and not requiring extensive laboratory equipment, they require a careful balance between dissociation of the glycoprotein bonds and retention of the 3D conformational properties of the analyte that might be the basis for the analyte’s detection (*20*). Additionally, the reagents can be expensive and might have a limited availability – these features can preclude rapid deployment and scale up of diagnostic operations in the case of a local or global health emergency, such as the COVID-19 pandemic.

In this study, we describe the SHEAR saliva collection device (SCD), a safe and simple Point-of-Care (POC) mechanical sample homogenization device made up of 3 main components: a soft foldable funnel, shearing filter and a collection tube. We report a reduction in viscosity with improvement in uniformity and increase in total protein concentration in saliva samples processed with SHEAR SCD compared to native, saliva processed by freeze-thawing, and supernatant. These outcomes were attributed to the mechanical shearing of saliva by the shearing filter. In addition, rheology results showed that the sample homogenization achieved by SHEAR SCD were comparable to conventional saliva processing methods, 1) freeze-thawing, 2) centrifugation, and 3) chemical homogenization (Dithiothreitol). Using rapid antigen testing (ART) with SARS-CoV-2 nucleocapsid protein, we demonstrate that processing of saliva with SHEAR SCD improved diagnostics performance. Furthermore, the wide and soft funnel conforms and fully covers the donor’s mouth reducing the risk of others being exposed to contagion during saliva donation. Our findings from backflow and food particle tests display that the unique shearing filter also limits the backflow of saliva and reduce the size and count of food particles, enhancing user safety due to prevention of spillage and ingestion of the content in the collection tube and mitigating potential interference with detection assay due to particle contaminant. Lastly, we also show SHEAR SCD is easy to use as assessed in a formal qualitative user study. These findings validate the functional capability and user friendliness of SHEAR SCD for the collection and processing of saliva samples especially in the POC setting.

## Results

### SHEAR SCD

The advent of the COVID-19 pandemic resulted in a sudden surge in demand for widespread SARS-CoV-2 testing and supply chain bottlenecks amplified the shortage of PPEs, medical equipment, and materials for diagnostic tests (e.g. nasopharyngeal swabs and reagent required for PCR). SHEAR SCD was designed in response to rigid funnel shortages for collecting saliva, and to address potential procedural bandwidth limitations with regards to sample processing, among other factors during the substantial ramp-up timeframe required to mobilize large-scale testing. SHEAR SCD is made of 3 main components: a soft foldable funnel, a shearing filter, and a collection tube (**Fig 1A**). Collection of saliva samples with SHEAR SCD involves 6 steps (**Fig 1B**) and 3 main activities: 1) deposition of saliva, 2) processing of saliva and, 3) funnel disposal. The usage of SHEAR SCD starts with pulling of the pull tabs to open the funnel and the deposition of saliva. Once enough saliva is collected in the funnel, the funnel is sealed and folded to press down and squeeze saliva through the shearing filter into the collection tube. Once enough saliva is collected in the collection tube, the funnel can be disconnected and disposed while the collection tube is closed with a screwcap. SHEAR SCD underwent product design cycles for an enhanced user experience and improved manufacturability. The high-fidelity prototypes, used for all testing in this study were manufactured through injection molding techniques under ISO13485 conditions. The early lab prototype (before the redesign process) is shown in **Fig 1C**. It was constructed within a week with readily available materials and technology. The filters were 3D printed using an inert and rigid photopolymer with a liquid resin printer. Piping bags commonly used for baking purposes were modified to construct the soft funnel and a hole was bored into a screwcap of a 15 ml centrifuge tube which houses the top filter module, and the centrifuge tube acted as the collection tube.

**FIG. 1.**
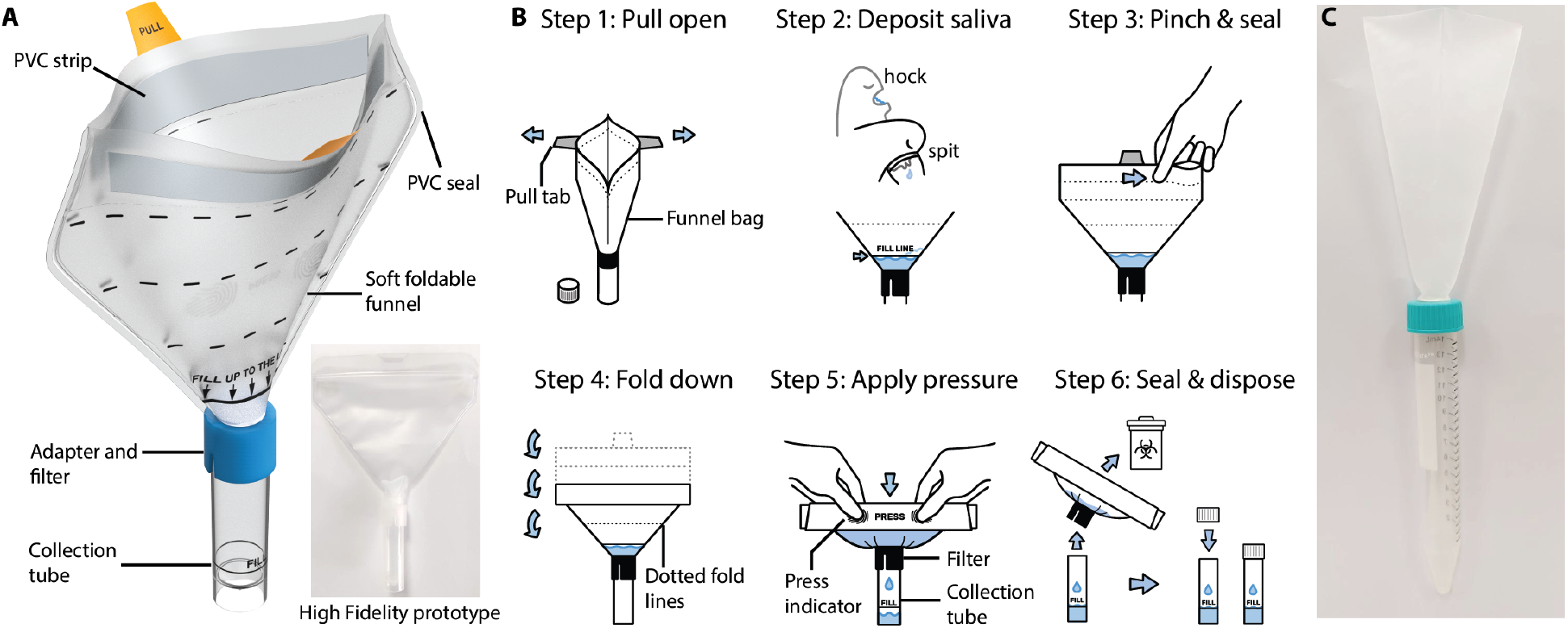
SHEAR SCD product design and instruction for use. (**A**). SHEAR SCD with labelled components. (**B**). Graphic of the instruction for use of SHEAR SCD. (**C**). Photo of the early lab prototype of SHEAR SCD.

### SHEAR SCD characteristics for saliva collection

The sample recovery test was conducted to investigate the sample retention in SHEAR SCD. When 2g saliva samples were pipetted directly to the base of the funnel, the average weight loss of saliva in SHEAR SCD and commercial, rigid SCD were 0.1710 ± 0.0227g and 0.0986 ± 0.0034g, respectively (**Fig 2A**). When saliva was pipetted to the middle of the SHEAR funnel the weight loss was 0.202 ± 0.0246g.

**FIG. 2.**
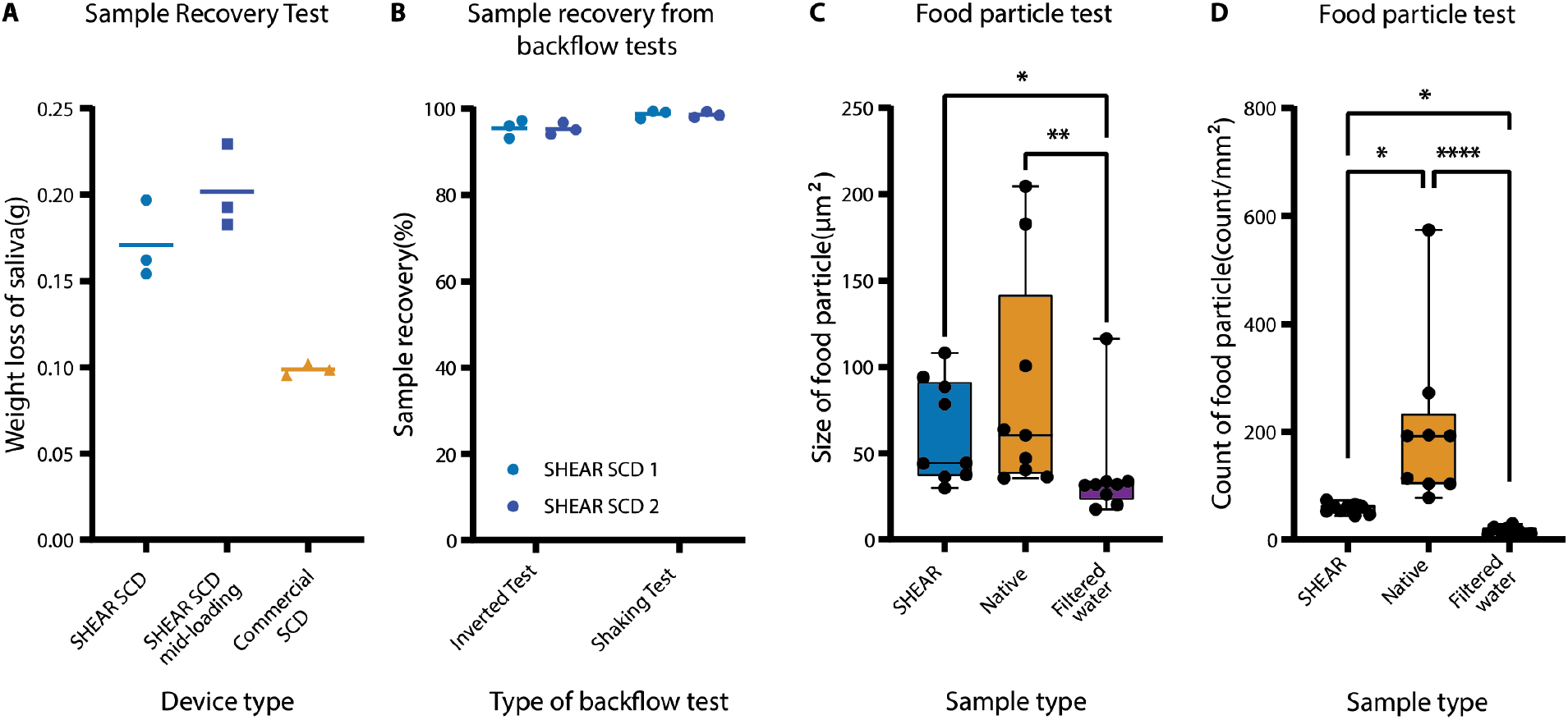
Sample recovery, backflow and food particle test results. (**A**). Weight loss of saliva from sample recovery testing of SHEAR SCD (n=3), SHEAR SCD with sample loading in the middle of the funnel (n=3), Commercial SCD (n=3). (**B**). Sample recovery of saliva samples from backflow tests; inverted test and shaking test, two SHEAR SCD devices (SHEAR SCD 1 and SHEAR SCD 2, N=3). (**C**). Size of food particles found in SHEAR (SHEAR SCD-processed sample), Native (Native sample) and Filtered water in the food particle test. (**D**). Count of food particles found in [count/mm^2^] of SHEAR (SHEAR SCD-processed sample), Native (Native sample) and Filtered water in the food particle test. N = number of replicates, n = number of devices. Lines in (**A-B**) represent the mean value. For (**C-D**) whiskers represent maximum and minimum values and the box represents the median value, 25th and 75th percentile. \*\*\*\**P*<0.0001,\*\*\**P*<0.001, \*\**P*<0.01 \**P*<0.05

POC tests commonly include a buffer in the collection tube for enhanced analyte interfacing. Improper handling of a kit during the saliva donation process may lead to accidental ingestion of buffer medium from the collection tube. To test if SHEAR SCD filter stops backflow of saliva (or saliva-buffer mixture) from the collection tube, we performed two sample recovery tests with saliva present in the collection tube: after shaking the tube with SHEAR SCD attached in a horizontal position, and after inverting the tube with SHEAR SCD attached upside-down. The average weight loss of saliva in the shaking test and the inverted test were 0.0268 ± 0.0151g and 0.0932 ± 0.0307g respectively, which corresponded to a sample recovery rate of 98.8% and 95.4% respectively (**Fig 2B**).

Presence of large food particles in saliva samples may possibly introduce interference into the downstream analytical tests. The food particle analysis was conducted to test the efficacy of the shearing filter in SHEAR SCD to trap particle and reduce contamination. The median size and count of food particles found in SHEAR SCD-processed sample were 44.38 μm^2^ (95% Confidence Interval (CI), 36.38 - 93.86) and 60.86 count/mm^2^ (95% CI, 46.07-64.84) respectively - a 36.1% reduction in size and a 215.8% significant reduction in count (*P*<0.05, Kruskal Wallis with Dunn’s test at α = 0.05) as compared to the native sample before processing (**Fig 2C-D**).

### SHEAR SCD enables saliva homogenization and analyte release

To assess the sample processing performance of SHEAR SCD, the viscosity of SHEAR SCD processed saliva samples was measured across a shear range of 50-3000s^−1^ and compared to saliva samples treated with other conventional saliva processing methods and an ART buffer solution. The saliva samples and ART buffer solution exhibited a non-Newtonian, shear thinning behavior; decreasing viscosity with increasing shear stress (**Fig 3A**). While no statistically significant differences in viscosity of saliva were detected between SHEAR SCD processed saliva and all measured samples (Kruskal Wallis with Dunn’s test at α = 0.05), at low shear rate (50s^−1^) the viscosity of SHEAR SCD processed saliva was lower (7.20 cP (95% CI, 5.02 - 9.28)) than Native, Freeze-Thawing and DTT processed saliva, but higher to that of Supernatant and ART buffer (**Fig 3B**). Except for DTT processed saliva, a similar trend of viscosity differences between SHEAR SCD processed and all measured samples was observed at high shear rate (3000s^−1^) (**Fig 3C)**. Additionally, we calculated the coefficient of variation (CV) of viscosity to assess the uniformity of the measured samples. Enhanced uniformity of viscosity was observed in SHEAR SCD processed saliva; CV of SHEAR SCD processed saliva was lower than all but the Supernatant samples (**Fig 3D**).

**FIG. 3.**
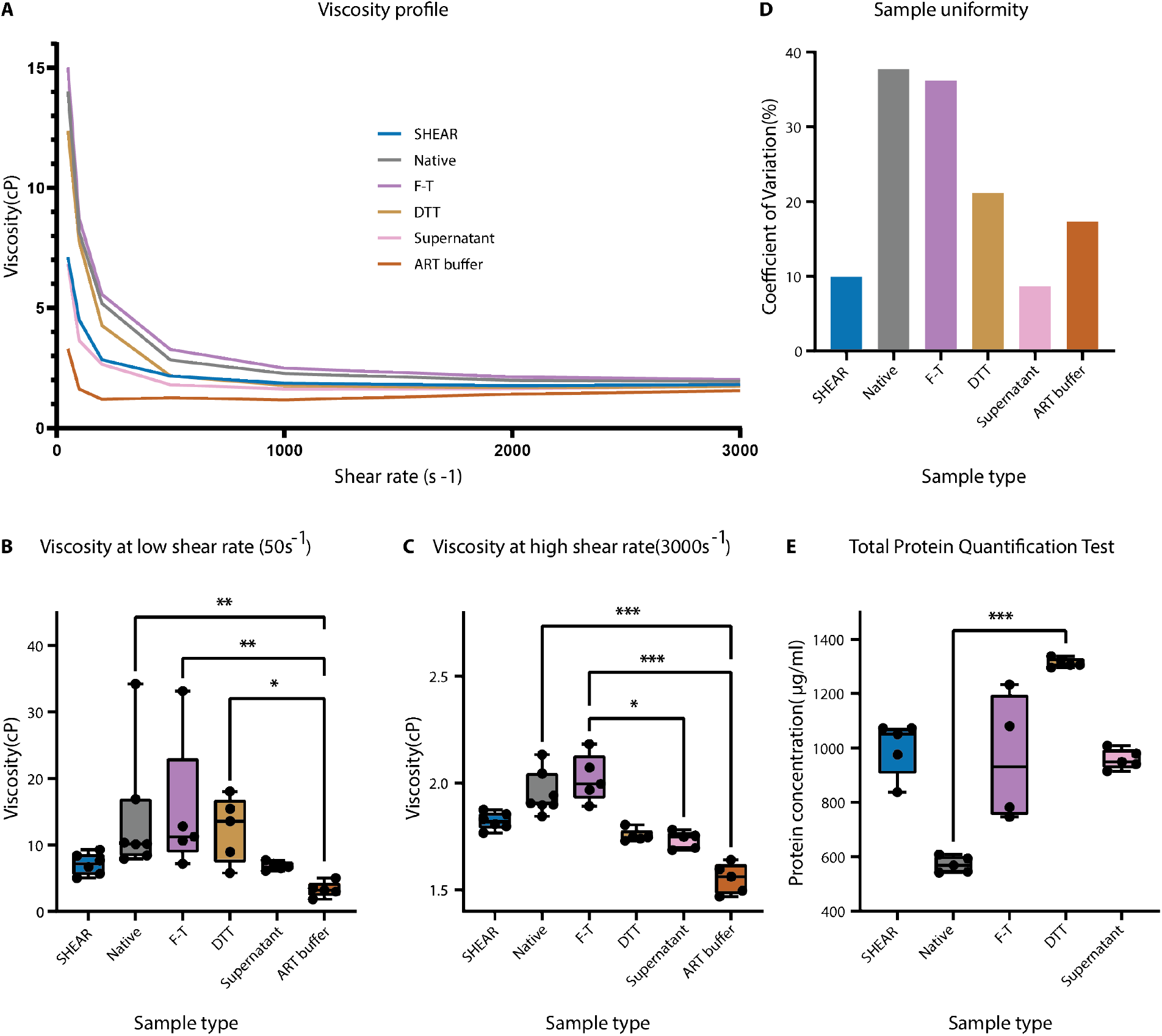
Viscosity and total protein concentration of saliva samples. (**A**). Viscosity profile of fluid samples measured with rheometer: SHEAR (SHEAR SCD processed saliva, N=6), Native (Native saliva, N=7), F-T (Freeze-Thawing processed saliva, N=5), DTT (DTT processed saliva, N=5), Supernatant (Centrifuged processed saliva, N=5) and ART Buffer (N=5) over a shear range from 50*s*^−1^ to 3000 *s*^−1^. (**B**). Viscosity of the fluid samples measured with rheometer at low shear rate: 50*s*^−1^. (**C**). Viscosity of the fluid samples measured with rheometer at high shear rate: 3000*s*^−1^. **(D**). Average sample uniformity of the samples measured with rheometer over the shear range from 50*s*^−1^ to 3000 *s*^−1^. (**E**). Estimation of total protein concentration of SHEAR (SHEAR SCD processed saliva, N=5), Native (Native saliva, N=5), F-T (Freeze-Thawing processed saliva, N=4), DTT (DTT processed saliva, N=5), Supernatant (Centrifuged processed saliva, N=5). Whiskers represent maximum and minimum values and the box represents the median value, 25th and 75th percentile. \*\*\**P*<0.001, \*\**P*<0.01 \**P*<0.05

A bicinchoninic acid (BCA) assay was conducted to assess and compare the total protein concentration of 5 saliva samples treated with different saliva processing techniques. The total protein concentration in SHEAR SCD processed saliva with 1051 µg/ml (95% CI, 837.7 - 1072.3) was higher when compared to Native, Freeze-Thawing and Supernatant but lower than DTT-processed saliva (**Fig 3E**). Except for DTT processed saliva (p<0.001), the increase was not statistically significant (Kruskal Wallis with Dunn’s test at α = 0.05).

### SHEAR SCD pre-processing affects the diagnostic performance of an ART

To investigate the effects of improved saliva’s biophysical properties on ART, paired saliva samples processed with SHEAR SCD and commercial SCD were spiked with SARS-CoV-2 nucleocapsid protein at the limit-of-detection (LOD) concentration level and tested with a commercial SARS-CoV-2 ART kit. Presence of a filtered out yellow residue was detected in the sample well of the ART kit for several individuals with less of that residue detected in the paired, SHEAR-preprocessed samples (**Supplementary Fig S1**). We measured the change in the intensity of the background after passing of the sample, and the relative intensities of the test and control lines after 20 and 30 mins from dropping the samples into the sample well of an ART cassette. While there was no statistically significant difference in the intensities of the background and the lines between the two SCD types (Wilcoxon signed-rank test at α = 0.05) at either of the timepoints, the samples processed with the commercial SCD led to formation of a visible test line for 6/10 individuals and SHEAR SCD improved that result by facilitating the test line formation in the samples from 9/10 of the same individuals (**Fig 4A**). No correlation was detected between the change in the background intensity and the relative intensity of the test line.

**FIG. 4.**
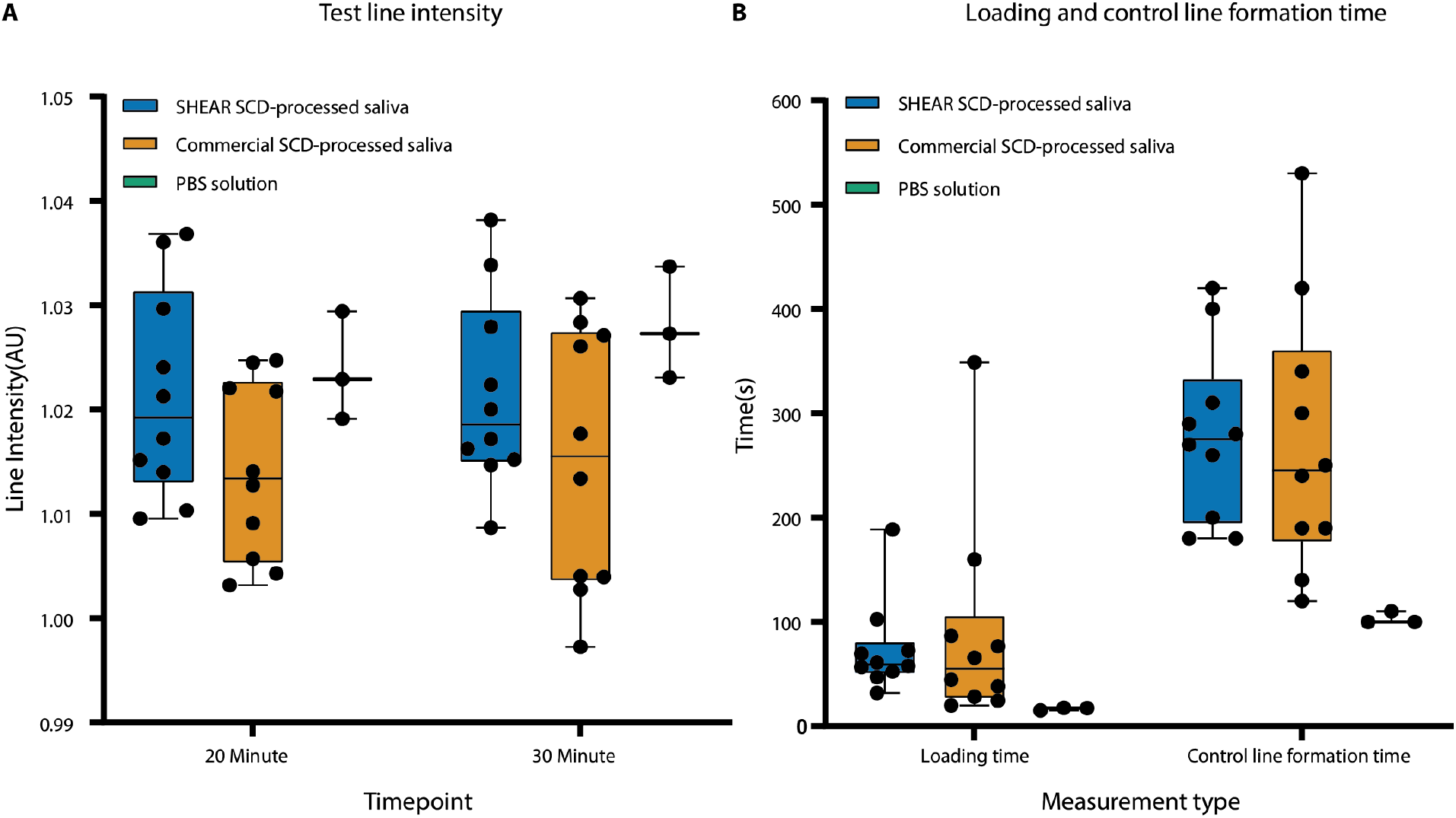
Test line intensity, loading time and control line formation time. **(A)**. Test line intensity for SHEAR SCD-processed saliva (N=10), Commercial SCD-processed saliva (N=10) and PBS solution (N=3) at 20-minute and 30-minute timepoints. **(B)**. Loading time and control line formation time of SHEAR SCD processed saliva (N=10), Commercial SCD processed saliva (N=10) and PBS solution (N=3). Whiskers represent maximum and minimum values and the box represents the median value, 25th and 75th percentile. No statistical difference detected with Wilcoxon signed-rank test at α = 0.05.

We additionally investigated the effects of SHEAR SCD on temporal dynamics of sample loading and line formation. There was no difference in the median loading time between the two groups (59.36 s vs 55.18 s, for SHEAR SCD and commercial SCD sample group, respectively; p = 0.96 Wilcoxon signed-rank test), however, SHEAR SCD sample preprocessing decreased the variation in the loading time by two times - with a CV of 0.75 vs 1.82 in the SHEAR SCD and commercial SCD test groups, respectively. For comparison, the median loading time of the control samples (PBS; N=3) was 17.5 s with CV of 0.08. We further assessed the time required for the sample after loading to pass the membrane assay across the result window and lead to the formation of the control line. The time required to form the control line was not significantly different between the two groups (3.25 min and CV of 0.42 vs. 3 min and CV of 0.31 in the SHEAR SCD and commercial SCD sample group, respectively) and was half of that in the control group (1.5 min with CV of 0.06) (**Fig 4B**). Lastly, no correlation was found between liquid migration speed and line intensity for test and control line (*R*^2^ = 0.035 and 0.095 respectively) (**Supplementary Fig S2**).

### SHEAR SCD is user friendly

The collection of saliva using SHEAR SCD includes an additional processing step to be done by the saliva donor - folding down the funnel to push the saliva sample through the filter into the collection tube. To investigate the amount of pressure required for users to push their saliva through the filter, the pressure generated within funnel during the processing step was measured. The maximum pressure generated within the funnel during the processing step was 7.50 ± 1.55 PSI.

15 healthy individuals participated in the user study of the SHEAR saliva collection device from February 2022-June 2022. There were eight female and seven male participants with a median age of 27 (range 23-60 years). Participants were provided with an instructional video (n1labs.org/shear-scd). Of note they did not require additional help throughout the user study and were able to complete the processing of saliva with SHEAR SCD independently. The participants were asked to donate saliva into a standard, commercial rigid funnel shortly after, within one session. All participants participated in a semi-structured interview which was conducted after the use of SHEAR SCD. 3 main themes emerged from the responses from the interviews and data saturation was achieved with 15 participants – no new themes emerged after participant 11.

#### Usability and functionality of SHEAR SCD

Participants commented positively towards SHEAR SCD and provided some suggestions for improvements. 93.3% of the participants responded favorably towards the usability of SHEAR SCD, specifically regarding the ease of use and straightforwardness of the device. Participant 6 shared, “*[The SHEAR device] is a really straightforward device and I don’t think it is complicated to use*.*”* Furthermore, all participants found that the instructions for use of SHEAR SCD were simple, clear and easy to understand which boosted their confidence in handling the device. According to Participant 6, *“The [instructional] video really helped. I wouldn’t have the confidence to use it if I have not watched the video*.*”* 13 out of 15 users commented positively on the additional functionality that SHEAR SCD funnel offers. The participants described an increased comfort level, ease of saliva donation and increased perceived safety level due to the large funnel size. In addition, two participants commented on the enhanced suitability of SHEAR SCD for elderly due to the large funnel size. Nevertheless, 11 participants highlighted the more simplistic nature of the commercially available SCD. For instance, Participant 10 shared, *“The [commercial device] was simpler, but the [SHEAR device] actually has a function*.*”* The difficulties reported by participants were mainly attributed to the presence of a push back of the saliva into the funnel during the squeezing and rolling steps, unclear saliva level indicator and insufficient air trapping within the funnel. Lastly, one participant highlighted the challenges users with disability might encounter during the processing step of SHEAR SCD.

#### Saliva as a biological material for diagnostic tests

All participants reported to have prior experience with diagnostic tests that involve collection of biological samples such as urine, blood and nasal swabbing. Four participants had undergone prior saliva-based diagnostic tests which involved either spit collection or/and cheek swabbing. In general, the participants were highly receptive towards saliva as a biological material for diagnostic tests and most had no concerns of processing of their own saliva. All participants commented positively on the ease of collection of saliva samples, which was attributed to comfortability (not painful nor invasive), simplicity of the collection process and high confidence for self-collection. While half of the participants highlighted their preference for saliva collection over nasal swabbing, some participants had reservations regarding the collection method. Five participants expressed concerns on the safety aspect of saliva collection, citing higher environmental contamination risk due to higher probability of saliva sample spillage during the discarding process when compared to nasal swabbing. In addition, two participant preferred cheek swabbing over spit saliva collection. Overall, the comments on saliva collection methods were largely positive suggesting the receptiveness and potential of saliva as a biological material for diagnostic applications (**Supplementary table 1**).

#### User’s consideration for the adoption of diagnostic kit with SHEAR SCD

When participants were asked to list their consideration for the adoption of a diagnostic kit that contains SHEAR SCD, they highlighted the accuracy of the diagnostic kit, cost, hygiene, safety and, ease-of-use as their main considerations for adoption. Furthermore, participants mentioned that the recognition status of saliva test by government, the amount of saliva required for accurate testing and the pre-testing requirement of no drinking, eating and brushing of teeth for a time period were some saliva-related concerns too (**Supplementary table 1**).

## Discussion

### SHEAR SCD scalably fulfils basic requirements for saliva collection

A saliva collection device is typically classified as a medical device. Hence, safety and effectiveness are two essential components that need to be demonstrated. While uncommon, several accidental exposures to buffer solutions from diagnostic test kits resulting in minor health outcome have been reported (*21*). In our backflow tests, 99% and 95% of the saliva sample was retained in the collection tube after the inverted and horizontal backflow tests respectively, demonstrating SHEAR filter’s ability to reduce backflow of the saliva samples and mitigate the risk of spillage leading to lowered incidence of such potential accidental exposure of saliva sample and buffer solution to the user. Furthermore, collection of saliva with SHEAR SCD provides an additional filtering function to reduce food debris preventing potential interference in diagnostic assays (*22*). In our food particle test, the 36.1% and 215.8% reduction in size and number of food particles observed in SHEAR-processed samples, potentially decrease the chance of particle interference with detection assays. Aside from particle contamination, saliva retention within the funnel itself can lead to a decrease in yield of the analyte of interest, potentially impairing accuracy of the diagnostic test. While, SHEAR SCD demonstrated a higher weight loss of saliva when compared to commercial SCD in the sample recovery test, the sample recovery rate was still above 90%, which is sufficient to perform follow up diagnostic assay.

### SHEAR SCD improves the physical properties of saliva for diagnostics analysis

Saliva is mainly produced by three pairs of salivary glands, the submandibular, parotid, and sublingual glands (*8*) and numerous minor glands in the mouth and throat. The composition and biophysical properties such as viscosity of saliva produced from each gland varies (*23*), hence the collection methods and types of saliva collected plays a key role in accurate detection of specific biomarkers. Saliva with high viscosity is challenging for both laboratory-based testing and POC diagnostic applications involving lateral flow assay and paper-based analytical devices. The observed reduction in saliva viscosity, due to the mechanical shearing effects of the SHEAR SCD filter, can ease the pipetting procedures in laboratory-based saliva testing reducing pipetting errors, risk of environmental cross contamination and processing time required (*10, 11, 13*). Furthermore, while the viscosity of supernatant after centrifugation was comparable to the viscosity of SHEAR SCD processed saliva, implementation of SHEAR SCD for POC application is more feasible since it does not require additional equipment. Freeze-Thawing is a commonly used pre-processing method for the preparation of saliva samples to reduce viscosity through the breaking down of mucopolysaccharide (*24*). However, no decrease in viscosity was observed in the Freeze-Thawing saliva samples in our viscosity testing. This could possibly be due to the absence of the centrifugation step that was present in studies that demonstrated reduction in viscosity which remove precipitates from thawed samples (*12, 25*).

Color formation with the BCA assay is mainly due to the number of peptide bonds and presence of specific amino acids (*26*). In our total protein concentration study involving the use of BCA assay, an increase in total protein concentration was observed in all processed saliva samples when compared to the native saliva samples. Breaking down mucopolysaccharide can potentially reduce the viscosity of the saliva sample and release analytes bound to the mucin networks. A previous study has attributed the increase in total protein concentration in mechanically processed (magnetically-beat) saliva samples to the disruption of mucin networks, which released protein for increased detection by BCA assay (*14*). Specific to COVID-19, a disruption of mucin networks can possibly release SARS-CoV-2 spike protein, which was recently found to bind to the sialic acid glycan end of mucins (*27*), and enhance the virus’ detection. Aside from DTT-processed saliva samples, the increase in protein concentration was comparable among samples processed with all other methods. DTT-processed saliva samples displayed the highest increase in protein concentration suggesting the greatest disruption of mucin networks. DTT is a commonly used reducing reagent for chemical homogenization of saliva and sputum that homogenizes samples through splitting of disulfide bond of proteins. While effective, sample homogenization with DTT or other harsh chemical reagents is not suitable for every application due to imposing alterations in biochemical properties of the analytes that may lead to their impaired detection with the sensitive detection methods, and even reduce concentration of certain biomarkers. For example, sample homogenization with DTT has been shown to reduce the concentration of e.g. sputum myeloperoxidase (*19, 28*), a potential biomarker for chronic obstructive pulmonary disease (COPD) management and monocyte chemoattractant protein-1 and macrophage inflammatory protein (*18, 20*), mediators associated with COPD.

### SHEAR SCD effects on ART testing

Saliva samples processed with SHEAR SCD demonstrated a mild improvement in the test line intensity. Interestingly, previous studies demonstrated an increase in signal intensity of test lines in samples with higher viscosity suggesting the amplification of the signal intensity could possibly be due lower sample flow rate which leads to increased incubation time of the analyte at the test lines (*29*) and elevated reaction time at the conjugate pad (*30*). Several other studies have corroborated the effects of viscosity on the flow rate of the liquid sample across the nitrocellulose strips of a lateral flow assay (*31, 32*). In contrast, in our study, no correlation was found between liquid migration speed and line intensity for the test and control lines. Instead, we hypothesize, that as the nucleocapsid protein was spiked into the liquid fraction of the saliva samples after passing through SHEAR SCD and the commercial SCD, it is plausible that the higher uniformity and lower viscosity of the SHEAR SCD-processed samples allowed better dispersion of the analyte and the buffer during the brief, 1 second-long vertexing, enhancing the inclusion of the analyte within the sample dropped onto the ART’s sample well. This may be indicative of the benefit of homogenizing the sample before its interfacing with the collection buffer and an improved analyte distribution for diagnostic of the analytes with varied presence in different fractions of the collected sample.

Of note, the test line intensity was generally low in this study due to the spiking of the nucleocapsid protein at the LOD concentration of the antigen to challenge the test to the extreme cases and reflect individuals with low viral load. The improved visibility of the test lines after sample pre-processing with SHEAR SCD could influence the judgment of the test result and potentially lead to a reduction in false negatives and a limitation of the intervention delays.

The sample well of an ART and the membrane assay allow only the liquid fraction to pass through to interact with the antibody conjugated strips to form the test and the control lines. Accordingly, SHEAR SCD had no effect on temporal dynamics of the sample migration after loading. However, SHEAR SCD lowered the variation in the sample loading time, which facilitate identification of a universal readout period for the most reliable results by lowering the interindividual, natural variation of saliva samples.

### SHEAR SCD offers benefits to user experience and safety

Developing an engineering solution for a medical application, benefits from including all stakeholders in the co-development process (*33*). Through interviews with the users, we sought to identify the attitudes towards SHEAR SCD, particularly as an additional step (folding the funnel) is expected from the saliva donor. In general, participants of the user study commented that SHEAR SCD was straightforward and easy to use, with simple and easy to understand instructions for use. The difficulties reported for SHEAR SCD were mainly attributed to the pushback of saliva during the squeezing and folding process, unclear saliva level indicator and insufficient air trapping within the funnel. Targeted refinements of SHEAR SCD based on the highlighted difficulties will benefit the usability and functionality for enhanced user-experience. In our pressure testing, the measured pressure required to squeeze saliva through the filter was 7.50 ± 1.55 PSI, a pressure that an average 70-year-old female can generate through pulp pinching (*34*), which was consistent to findings from user study where no difficulties in squeezing of saliva due to pressure required were reported.

Transmission of respiratory viruses can occur through direct contact with aerosol and droplets containing virions (*35*). It has been found that sneezing and coughing forcefully expels droplets into the surrounding environment (*36*) hence spitting and hocking actions during saliva collection could potentially lead to release of such aerosols and droplets increasing the risk of transmission of such diseases. Furthermore, in a recent study, an incidence rate of 27.8% of saliva contamination on exterior surface of collection tube was reported (*37*). Saliva collection with SHEAR SCD covers the user’s mouth and nose during the donation process, containing the dispersion of droplets within the funnel and prevents saliva leakage to the exterior surfaces of the device. In addition, the foldable funnel enables the closing of the opening which leads to a reduction in the exposure time between donated saliva and the environment. While nasal swabbing methods do not lead to release of such droplets, three participants from our user study reported sneezing after the conduct of nasal swab, which is a drawback of nasal swabbing for detection of airborne respiratory viral infections in a mass testing setting. One participant in our study reported history of epistaxis after nasopharyngeal swabbing perfomed by a healthcare worker. While nasal swabbing is a less invasive procedure, there is still a risk of epistaxis. In addition, a participant expressed concerns of nasal swabbing of children and elderly due to underdevelopment of skull and lower tissue integrity, respectively. Compared to nasal swabbing, ease of saliva collection in terms of comfort and easier visual quantification of sample volume facilitates the extraction of the required amount of sample for a follow up diagnostic test. In fact, a study revealed that 16% of their participant were fearful to conduct self-nasal swabbing leading to the occurrence of “fairly shallow swab” in some cases (*38*), which potentially amplifies the probability of false-negative in samples from those participants. The simplicity of saliva collection, coupled with the use of SHEAR SCD that brings increased comfort level and ease of saliva donation as commented by participants in our user study, points to SHEAR SCD’s potential for home-based and remote testing.

### Limitation

While offering multiple benefits, SHEAR SCD is not without limitations. The effect of mechanical shearing is limited, and even though no statistical significance was detected, the viscosity of SHEAR SCD-processed saliva was higher than that of buffer solution used in the ART. Additionally, in our study, while the participants were instructed to donate back throat saliva, drool saliva secreted from parotid gland were inevitably donated in the process, which resulted in the increased variability of viscosity within the individual’s saliva sample. Furthermore, in the total protein and viscosity experiments saliva samples collected from multiple participants were pooled, as both experiments required more amount of saliva than what an individual can donate in a sitting. Both of those factors may have contributed to increased variation in the saliva viscosity. To lower its effects, saliva samples in all groups were gently vortexed for 1 second right before each test.

The ART kit testing was used off label - the saliva sample were not the validated specimen listed in the instruction for use (IFU). Also, spiking of SARS-CoV-2 nucleocapsid proteins into processed healthy saliva samples limits the evaluation of the effects of mechanical shearing on mucin networks, which can potentially release SARS-CoV-2 virions trapped in that network. The benefits of SHEAR SCD for POC kits will require further testing in clinical trials with this study setting a foundation for such endeavour. Lastly, in the user study the mean age of the participants was 34 (ranging from 23-60 years) and only healthy individuals were recruited, hence findings from the study maybe have limited generalizability to elderly, children and people with disabilities. User study with participants representing wider age group and health status may capture a more comprehensive sentiment for SHEAR SCD among the real-world users, however, the results of our study demonstrate general acceptability of the device.

### SHEAR SCD’s relevance for saliva-based clinical testing beyond COVID-19

Saliva has an advantage over other diagnostic samples due to the easy, safe, and non-invasive nature of attaining the sample. Similar to the responses recorded from participants in our user study, patients also cited these among reasons for which they prefer oral fluid sampling when given the choice (*39*). The non-invasive nature of saliva collection may be greatly beneficial for broad deployment under epidemic and pandemic circumstances, but also in a routine care, especially for patients that requires inpatient and/or outpatient longitudinal tests. These factors may potentially increase compliance with the testing regime, lowering the stress exerted on patients, and potentially enabling remote monitoring.

As demonstrated in COVID-19 diagnostics, POC testing, including saliva-based, has a potential for an early detection of the infection, which in turn allows earlier management options to be commenced and a greater likelihood for successful therapy. The benefits extend to other infectious diseases – e.g. saliva-based POC tests can be used for the diagnosis of HIV (*40*) and Hepatitis C, where the presence of immunoglobulins can inform about the stage of the infection (*41*). Other infectious diseases, e.g. Hepatitis B, mumps and rabies, also has a potential to be detected in saliva (*42*). Beyond infectious diseases, salivary diagnostics are clinically applicable to monitoring drug abuse, hormone levels and a range of disease markers. For example, saliva-based monitoring of C-reactive protein (CRP), a non-specific inflammatory response factor, has a potential to assist in detection and tracking of inflammation (*43, 44*), enabling earlier detection of infection and supporting rapid decision making. Similarly, being able to monitor drug levels with saliva-based POC testing will help reduce the number of invasive blood tests (*45*).

While salivary diagnostics hold potential, they are not often used as first line tests or stand-alone tests. Broadly deployed applications of saliva-based POC testing are also not common. The challenges stem from minute quantities of the markers and sample variability (*46*). Improving the biophysical properties of saliva sample for diagnostics at the collection stage with SHEAR SCD, offers a range of benefits and may potentially contribute to the broader realization of saliva’s potential as a clinical sample beyond COVID-19, especially in POC applications.

In summary, we presented a novel saliva collection device with sample homogenization capabilities that enhances interface for downstream analytical processes. SHEAR SCD-processed saliva samples exhibited lowered viscosity with enhanced uniformity, increased total protein concentration and an augmented visibility of test line when analyte was spiked at LOD concentration and detected with an ART. Importantly, participants from the user study highlighted the ease of use of SHEAR SCD and preference of saliva collection over collection of other diagnostic samples. In sum, the strategic implementation of SHEAR SCD may play an important role in enabling rapid and self-contained of saliva processing for diagnostic applications, especially in the context of POC testing during outbreak, epidemic and pandemic conditions.

## Materials and Methods

### Manufacturing method of SHEAR SCD

SHEAR SCD is made out of three main components: a soft, foldable funnel, shearing filter with small pores housed in an adapter and a collection tube. The components were fabricated and assembled by an ISO 13485:2016 certified manufacturer (Forefront Medical Technology, Singapore). The soft funnels were made from die-cutting and heat sealing of PolyVinyl Chloride (PVC) sheets with the aid of the 2D computer aided diagram drawn with Solidworks (Dassault Systèmes, MA, USA). The filter was manufactured with micro-injection molding of Polypropylene (PP, K1P38AE) using MicroPower15t machine (Wittmann Battenfeld, Austria) and the adapters were constructed with injection molding of Polypropylene (PP, K1P38AE). Medical grade adhesive (Loctite 3921, USA) was used to bond the filter and the soft funnel to the adapter. Standard collection tubes were used.

### Saliva collection method and IRB

Saliva samples were collected from **24** healthy volunteers aged between **21** and **65** years in accordance with protocol approved by the Institutional Review Board of National University of Singapore (NUS-IRB-2021-15); written informed consent were obtained from each volunteer. Body temperature of each volunteer was taken prior to the study and the volunteers that exhibited acute respiratory infection symptoms were excluded. Volunteers were advised to avoid brushing of teeth and consumption of food and beverages for at least an hour before the collection of the saliva. The volunteers were advised to donate deep throat saliva. Each volunteer donated approximately 3ml of the sample. All samples were collected between 0900-1100 and 1300-1400.

### Pressure measurement

The maximum pressure generated within the funnel of SHEAR SCD during the folding and squeezing process was measured with a pressure transducer (Analog Pressure Sensor, Gravity) inserted at the bottom of the funnel. 1ml of saliva from three participants were added in each replicate test.

### Filter performance experiments

#### Food particulate test

In the food particulate test, 5 ml of filtered water (Mili-Q IQ 7000) was spiked with 0.1g of chili flakes and added to the funnel of one SHEAR SCD and one common, commercially available SCD (MicroCollect™ Saliva Collection Device, CD Genomics, USA). The saliva collected in the collection tube of each SCD, was added onto a glass slide and imaged at nine fixed points with 4× objective with a light microscope (ECLIPSE Ti-S, Nikon, Japan). Captured images were processed with ImageJ (National Institutes of Health, USA) for particulate counting and size measurement (**Supplementary Fig S4**).

#### Characterization of backflow

Two tests were conducted to investigate the amount of backflow of the saliva sample across the filter, from the tube to the funnel. In both tests, 2g of saliva were added into each SCD. In the first test, two SHEAR SCDs were placed on an orbital shaker (Orbital shaker OS-20, Boaco, Germany) and shook at 100 rpm for 60 minutes. In the second test, two SHEAR SCDs were vertically inverted and placed on a retort stand for 60 minutes. The tests were repeated in triplicates. The weight of the SCDs with and without the sample, as well as the weight of the collection tube before and after each test were recorded and used to calculate sample recovery for the evaluation of backflow.

#### Sample recovery

Sample recovery rate of SHEAR SCD and commercially available SCD (MicroCollect™ Saliva Collection Device, CD Genomics, USA) were measured and compared. 2g of saliva were added and processed in each SCD. The weight of the SCD with and without the sample, as well as the weight of the collection tube before and after sample processing for each replicate were recorded and used to calculate saliva sample retention and subsequently sample recovery rate. The experiment was repeated in triplicate.

#### Preparation of saliva samples for Total protein concentration and Rheology test

15 ml of saliva samples collected from eight healthy individuals were pooled and aliquoted into 5 parts of 3ml. Each aliquot was treated with different saliva processing technique: 1) Non-processed native samples, 2) SHEAR SCD-processed sample, 3) Freeze-Thawing-processed samples, 4) DTT-processed sample, and 5) Supernatant (centrifuged samples). For the Freeze-Thawing-processed saliva, the samples were first frozen in -20°C for an hour and subsequently thawed in room temperature for 30 minutes. DTT (Invitrogen, USA) were reconstituted in ultrapure water to a concentration of 10mM. The DTT-processed saliva was treated with 3mL of 10mM DTT. Lastly, the centrifuged saliva sample was obtained by centrifuging saliva in a bench top centrifuge machine (Centrifuge 5810, Eppendorf, Germany) at 3000g for 10 minutes and the supernatant was subsequently extracted and used in the measurements.

#### Total protein concentration test

The Pierce BCA Protein Assay Kit was purchased from Thermo Fisher Scientific (MA, USA) and used to estimate the total protein concentration of saliva samples. 1ml of each type of processed saliva samples (Native, SHEAR SCD-processed, DTT-processed, Freeze-Thawing-processed and Supernatant) were extracted and diluted with 2ml of ultrapure water. Standards and working reagent (WR) were prepared according to the user manual (*26*). 25ul of each standard and type of processed saliva samples (N=5) were pipetted onto the 96 well plate and 200ul of WR were added into each well. The samples were then incubated at 37 °C for 30 minutes and the sample colour intensity was measured using a microplate reader (Spark 10m, Tecan, Switzerland) at 562nm wavelength. The total protein concentration of the processed saliva sample was calculated in reference to the bovine serum albumin (BSA) standards. Protein concentration of one Freeze-Thawing processed saliva was above the working range of BCA assay and removed as an outlier.

#### Rheology

Viscosity of processed saliva samples (Native, SHEAR SCD-processed, DTT-processed, Freeze-Thawing-processed and Supernatant) were measured and analyzed. Processed saliva samples were mounted onto the Peltier plate of the rheometer (MCR302, Antoon Paar, Austria) and viscosity of the samples were recorded over a shear rate range from 50*s*^−1^ to 3000*s*^−1^ at 25°C using the cone plate measuring system (CP25-2). The measurement for each sample type was repeated 5-7 times.

### Rapid Antigen Testing procedures

*Panbio™ COVID-19 Ag Test Kit* (Abbott, USA) was used as the ART kit. Saliva samples of each volunteer were aliquoted into two parts. 500µl of saliva was extracted from each aliquot and processed with SHEAR SCD and commercial SCD (MicroCollect™ Saliva Collection Device, CD Genomics, USA). 1µg/ml SARS-CoV-2 nucleocapsid protein solution were prepared by diluting the 9.5 mg/mL stock solution (*47*). 4.5µl of SARS-CoV-2 nucleocapsid protein (1µg/µl) and 95.5ul of *Panbio* buffer solution were added into each processed saliva sample and a control solution - phosphate buffer saline solution (PBS). The mixture was vortexed, and 5 drops of the mixture were loaded onto the sample well of the ART cassette. The liquid mixture was allowed to migrate along the result window for 30 minutes. 30 minutes time-lapse videos (time-lapse interval = 10 seconds) of the development of each cassette was recorded with GoPro Hero 4 (GoPro, USA) under a custom-made lightbox to provide a fixed imaging condition (**Fig 5A**). The time-lapse videos were aligned and converted into 8bit image stacks with ImageJ (National Institute of Health, USA) for quantification of signal intensity (**Fig 5B**). The average pixel intensity of the regions of interest at the test line (*I*_*test line*_) and background (*I*_*background*_) were measured and test line intensity ratio, *I*_*test line*_ */ I*_*background*_, was calculated to reduce background noise (*48*) at 20 (t_20_) and 30 (t_30_) minutes after the sample loading. Test line formation was analysed through the measurement of the average pixel intensity of the region of interest at the test line and 20 pixels before and after test line (*I*_*test line*_, *I*_*before test line*_, *I*_*after test line*_ respectively). Test line is determined to be formed if *I*_*before* test line_ / *I*_test line_ and *I*_*after* test line_ / *I*_test line_ and average pixel intensity of *I*_*before* test line_ and *I*_*after* test line_ is not more than 120 arbitrary units (**Fig 5C**). Equations used for the ART procedures are shown in **Table 1**.

**FIG. 5.**
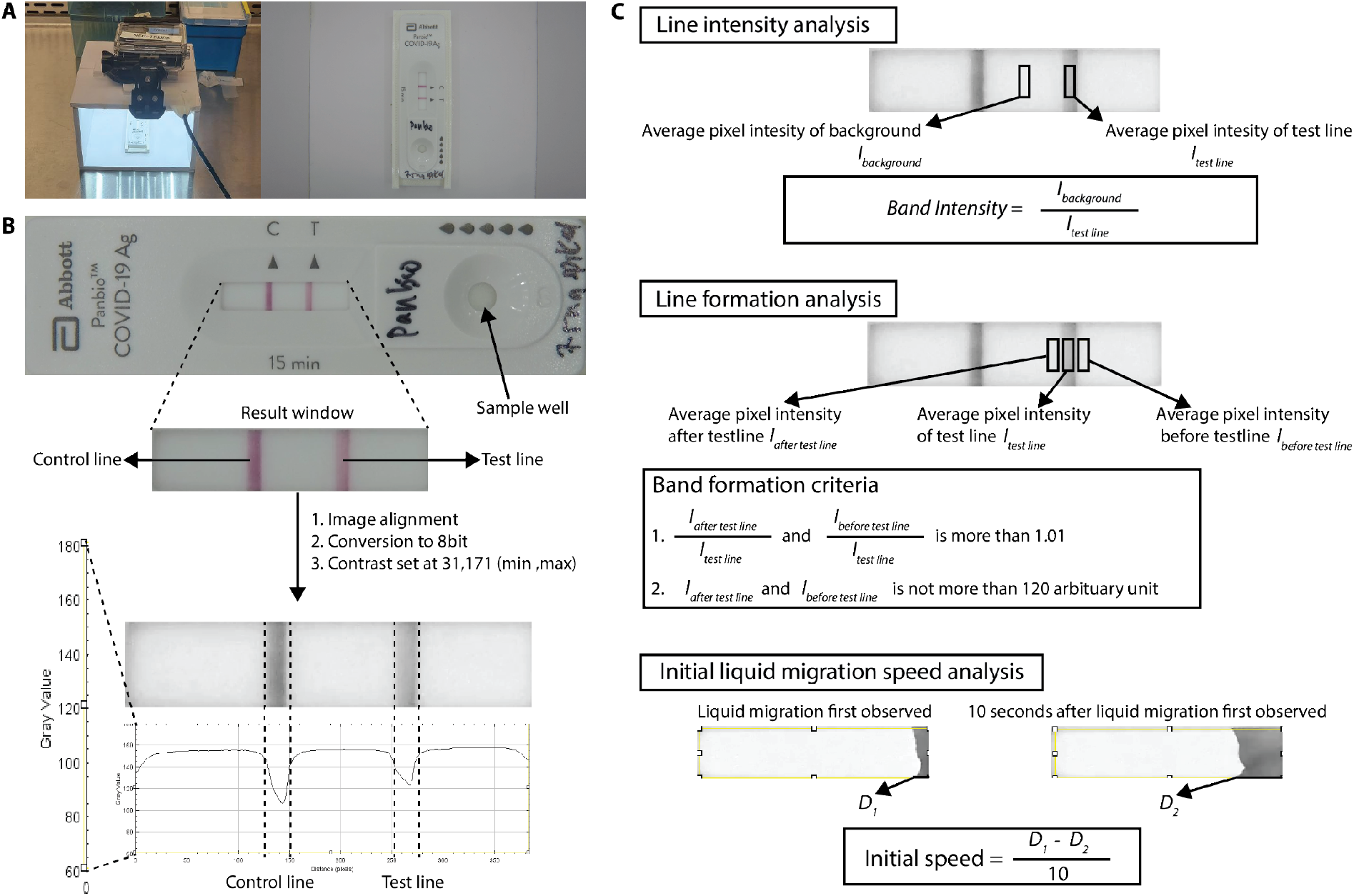
Analysis method for antigen rapid test. (**A**) Photo of the set-up for the capturing of time-lapse video for the antigen test experiment. (**B**) Antigen test cassette and general image processing step. (**C**) Description of analysis conducted in the antigen test experiment.

**Table 1.**
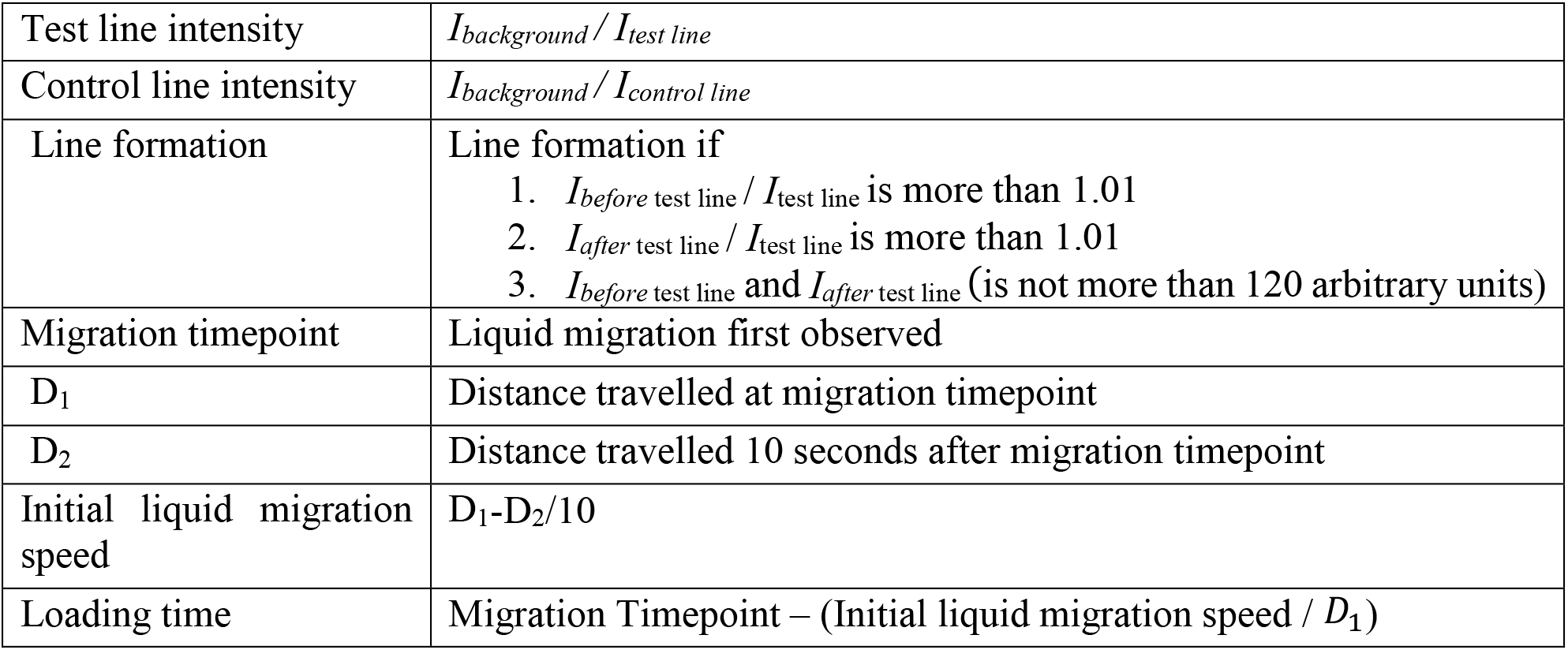
Equations and terminology in the analysis of Rapid antigen test experiment.

|  |  |
| --- | --- |
| Test line intensity | $I_{background} / I_{test\ line}$ |
| Control line intensity | $I_{background} / I_{control\ line}$ |
| Line formation | Line formation if<br>1. $I_{before\ test\ line} / I_{test\ line}$ is more than 1.01<br>2. $I_{after\ test\ line} / I_{test\ line}$ is more than 1.01<br>3. $I_{before\ test\ line}$ and $I_{after\ test\ line}$ (is not more than 120 arbitrary units) |
| Migration timepoint | Liquid migration first observed |
| $D_1$ | Distance travelled at migration timepoint |
| $D_2$ | Distance travelled 10 seconds after migration timepoint |
| Initial liquid migration speed | $D_1 - D_2 / 10$ |
| Loading time | Migration Timepoint – (Initial liquid migration speed / $D_1$) |

### User Study

The user study sought to evaluate the end user’s experience of the SHEAR SCD and a commercial SCD (MicroCollect™ Saliva Collection Device, CD Genomics, USA). Participants were asked to donate 1 ml of saliva to each of the two SCDs, and they were subsequently interviewed based on a semi-structured interview guide (**Table 2**). The average length of the interview was 21 minutes (range: 18-25 minutes). Participants’ responses were audio-recorded, transcribed verbatim and analyzed using thematic analysis, where the responses were descriptively labelled for primary coding. The labels were then categorized into different groups which were subsequently used to create broader themes.

**Table 2.**
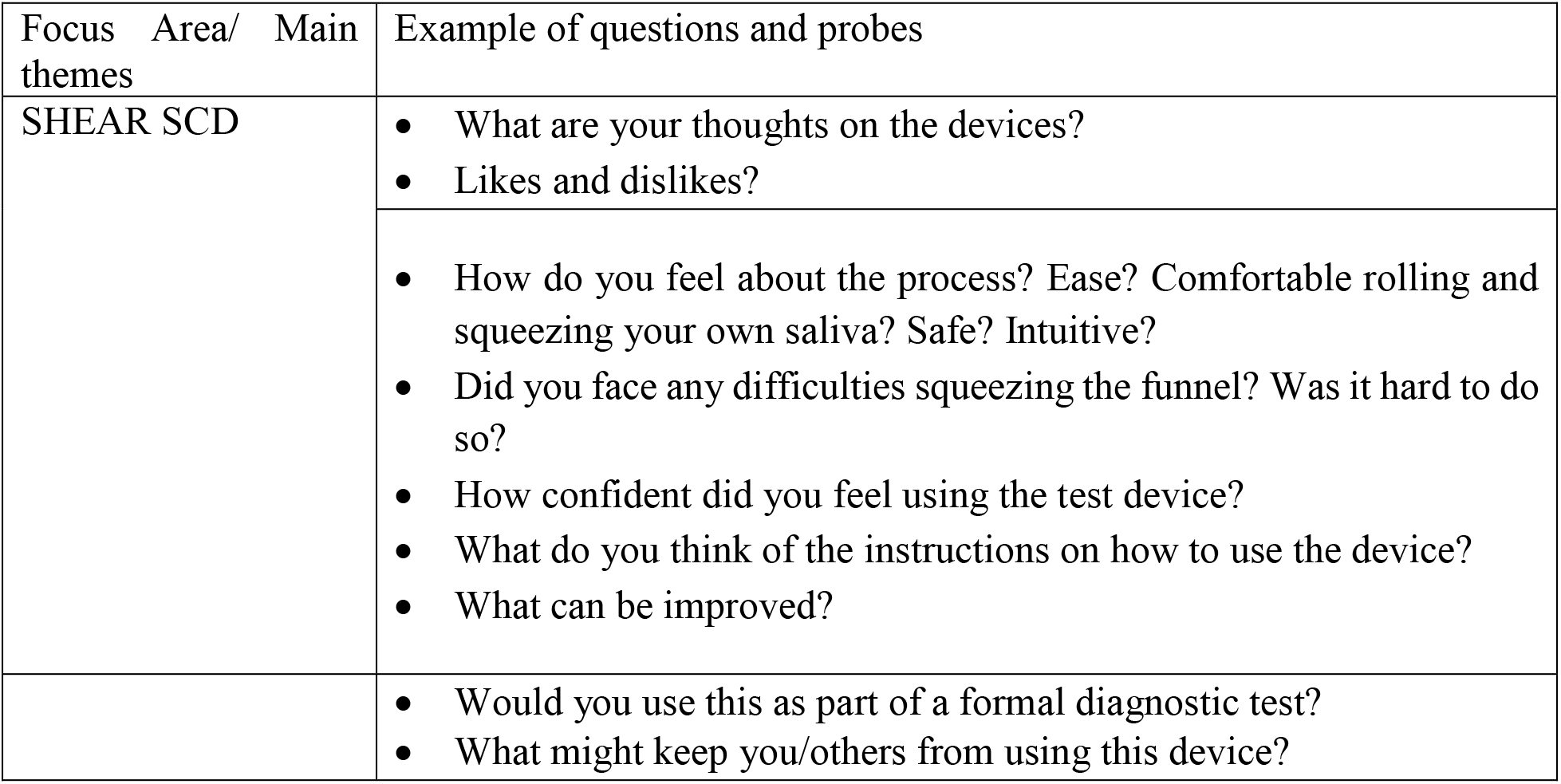
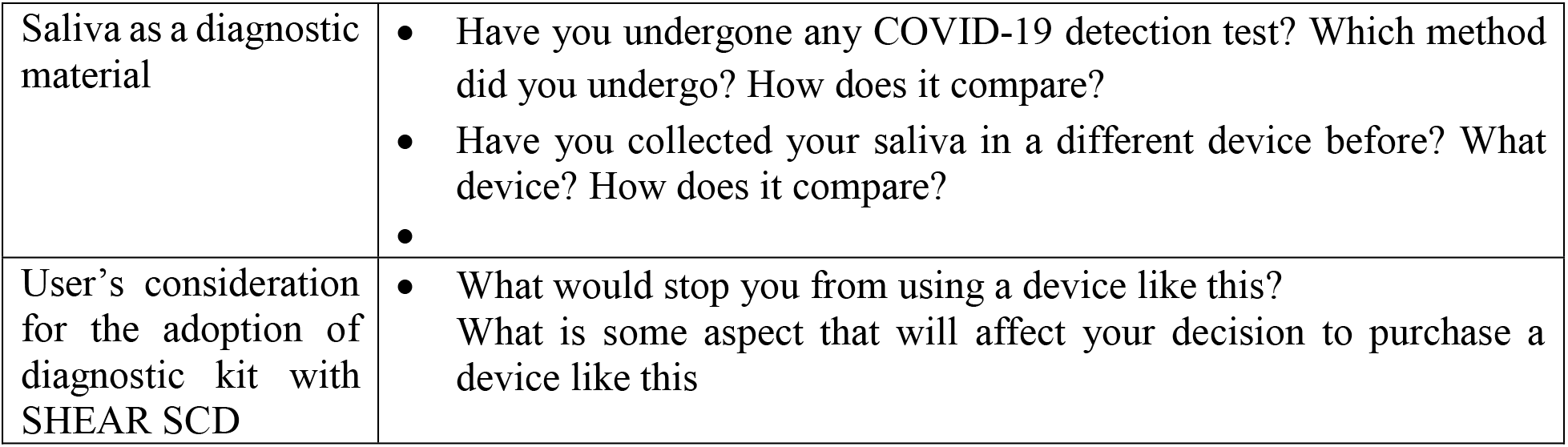
List of questions included in the usability study

| Focus Area/ Main themes | Example of questions and probes |
| --- | --- |
| SHEAR SCD | • What are your thoughts on the devices?<br>• Likes and dislikes? |
|  | • How do you feel about the process? Ease? Comfortable rolling and squeezing your own saliva? Safe? Intuitive?<br>• Did you face any difficulties squeezing the funnel? Was it hard to do so?<br>• How confident did you feel using the test device?<br>• What do you think of the instructions on how to use the device?<br>• What can be improved? |
|  | • Would you use this as part of a formal diagnostic test?<br>• What might keep you/others from using this device? |
| Saliva as a diagnostic material | • Have you undergone any COVID-19 detection test? Which method did you undergo? How does it compare?<br>• Have you collected your saliva in a different device before? What device? How does it compare?<br>• |
| User's consideration for the adoption of diagnostic kit with SHEAR SCD | • What would stop you from using a device like this?<br>What is some aspect that will affect your decision to purchase a device like this |

### Statistical Analyses

Statistical analyses were carried out using GraphPad PRISM 9 (version 9.3.1, GraphPad) Statistical significance was determined using the Kruskal Wallis test with Dunn’s test at alpha = 0.05 for the food particle, rheology and protein test, and Wilcoxon signed-rank test at alpha = 0.05 for the examination with ART.

## Supporting information

SHEAR SCD supplementary material

## Data Availability

All data needed to evaluate the conclusions in the paper are present in the paper and/or the Supplementary Materials.

## List of Abbreviation

ART: Antigen Rapid Test
BCA: Bicinchoninic acid
CV: Coefficient of Variation
DTT: Dithiothreitol
LOD: Limit of Detection
POC: Point of Care
SCD: Saliva Collection Device

## Acknowledgments

We thank members from Forefront Medical and Stuck Design for their active involvement in the design and optimization of the high-fidelity prototype of SHEAR saliva collection device. We thank C.R.He and A.Chia from the Industry Liaison office of National University of Singapore for their generous advice.

## Author contributions

Conceptualization: AB, PAM, DH

Methodology: AB, SWS, PAM, DH, VVL, RG, XLT

Investigation: AB, SWS, VVL, YS, NJ

Supervision: AB, PAM, DMA, JWEL, DH

Writing—original draft: AB, SWS, NJ, PAM, DH

Writing—review & editing: AB, SWS, VVL, RG, GY, DMA, PAM, DH

## References

1. U.S. Food and Drug Adminstration, In Vitro Diagnostics EUAs - Molecular Diagnostic Tests for SARS-CoV-2. (2022).

2. Austrialian Government Department of Health, COVID-19 test kits included in the ARTG for legal supply in Australia. (2022).

3. Singapore General Hospital, Saliva testing available in Singapore for Covid-19 pre-departure tests of travellers. (2021).

4. I. M. Ott, M. S. Strine, A. E. Watkins, M. Boot, C. C. Kalinich, C. A. Harden, C. B. F. Vogels, A. Casanovas-Massana, A. J. Moore, M. C. Muenker, M. Nakahata, M. Tokuyama Nelson, J. Fournier, S. Bermejo, M. Campbell, R. Datta, C. S. Dela Cruz, S. F. Farhadian, A. I. Ko, A. Iwasaki, N. D. Grubaugh, C. B. Wilen, A. L. Wyllie, I. R. t. Yale, Stability of SARS-CoV-2 RNA in Nonsupplemented Saliva. Emerg Infect Dis 27, 1146–1150 (2021).

5. S. B. Griesemer, G. V. Slyke, D. Ehrbar, K. Strle, T. Yildirim, D. A. Centurioni, A. C. Walsh, A. K. Chang, M. J. Waxman, K. S. George, A. J. McAdam, Evaluation of Specimen Types and Saliva Stabilization Solutions for SARS-CoV-2 Testing. Journal of Clinical Microbiology 59, e01418–01420 (2021).

6. G. Marais, N.-y. Hsiao, A. Iranzadeh, D. Doolabh, A. Enoch, C.-y. Chu, C. Williamson Brink, D. Hardie, Saliva swabs are the preferred sample for Omicron detection. medRxiv, 2021.2012.2022.21268246 (2021).

7. T. Pfaffe, J. Cooper-White, P. Beyerlein, K. Kostner, C. Punyadeera, Diagnostic Potential of Saliva: Current State and Future Applications. Clinical Chemistry 57, 675–687 (2011).

8. G. B. Proctor, The physiology of salivary secretion. Periodontol 2000 70, 11–25 (2016).

9. P. Ramachandran, P. Boontheung, E. Pang, W. Yan, D. T. Wong, J. A. Loo, Comparison of N-linked Glycoproteins in Human Whole Saliva, Parotid, Submandibular, and Sublingual Glandular Secretions Identified using Hydrazide Chemistry and Mass Spectrometry. Clinical Proteomics 4, 80–104 (2008).

10. F. Fronza, N. Groff, A. Martinelli, B. Z. Passerini, N. Rensi, I. Cortelletti, N. Vivori, V. Adami, A. Helander, S. Bridi, M. Pancher, V. Greco, S. I. Garritano, E. Piffer, L. Stefani, V. De Sanctis, R. Bertorelli, S. Pancheri, L. Collini, E. Dassi, A. Quattrone, M. R. Capobianchi, G. Icardi, G. Poli, P. Caciagli, A. Ferro, M. Pizzato, A Community Study of SARS-CoV-2 Detection by RT-PCR in Saliva: A Reliable and Effective Method. Viruses 14, (2022).

11. S. H. Tan, O. Allicock, M. Armstrong-Hough, A. L. Wyllie, Saliva as a gold-standard sample for SARS-CoV-2 detection. Lancet Respir Med 9, 562–564 (2021).

12. J. Noiphung, M. P. Nguyen, C. Punyadeera, Y. Wan, W. Laiwattanapaisal, C. S. Henry, Development of Paper-Based Analytical Devices for Minimizing the Viscosity Effect in Human Saliva. Theranostics 8, 3797–3807 (2018).

13. M. L. Landry, J. Criscuolo, D. R. Peaper, Challenges in use of saliva for detection of SARS CoV-2 RNA in symptomatic outpatients. J Clin Virol 130, 104567 (2020).

14. B. Johannsen, L. Müller, D. Baumgartner, L. Karkossa, S. M. Früh, N. Bostanci, M. Karpíšek, R. Zengerle, N. Paust, K. Mitsakakis, Automated Pre-Analytic Processing of Whole Saliva Using Magnet-Beating for Point-of-Care Protein Biomarker Analysis. Micromachines (Basel) 10, (2019).

15. S. Senapati, S. Das, S. K. Batra, Mucin-interacting proteins: from function to therapeutics. Trends Biochem Sci 35, 236–245 (2010).

16. G. Bilancio, P. Cavallo, C. Lombardi, E. Guarino, V. Cozza, F. Giordano, G. Palladino, M. Cirillo, Saliva for assessing creatinine, uric acid, and potassium in nephropathic patients. BMC Nephrol 20, 242–242 (2019).

17. Angel C. Y. Yu, Liam J. Worrall, Natalie C. J. Strynadka, Structural Insight into the Bacterial Mucinase StcE Essential to Adhesion and Immune Evasion during Enterohemorrhagic <em>E. coli</em> Infection. Structure 20, 707–717 (2012).

18. F. Wang, B. He, The effect of dithiothreitol on chemotactic factors in induced sputum of chronic obstructive pulmonary disease patients. Respiration 78, 217–222 (2009).

19. E. Grebski, C. Peterson, T. C. Medici, Effect of physical and chemical methods of homogenization on inflammatory mediators in sputum of asthma patients. Chest 119, 1521–1525 (2001).

20. M. M. K. Leader of the Working Group, G. Members of the Working, V. Keatings, R. Leigh, C. Peterson, J. Shute, P. Venge, R. Djukanović, Analysis of fluidphase mediators. European Respiratory Journal 20, 24s (2002).

21. The Government of Canada, Rapid antigen test kits and potential exposure to hazardous substances. (2022).

22. Z. Bai, H. Wei, X. Yang, Y. Zhu, Y. Peng, J. Yang, C. Wang, Z. Rong, S. Wang, Rapid Enrichment and Ultrasensitive Detection of Influenza A Virus in Human Specimen using Magnetic Quantum Dot Nanobeads Based Test Strips. Sens Actuators B Chem 325, 128780–128780 (2020).

23. G. Iorgulescu, Saliva between normal and pathological. Important factors in determining systemic and oral health. J Med Life 2, 303–307 (2009).

24. R. Mohamed, J. L. Campbell, J. Cooper-White, G. Dimeski, C. Punyadeera, The impact of saliva collection and processing methods on CRP, IgE, and Myoglobin immunoassays. Clin Transl Med 1, 19 (2012).

25. L. H. Schneyer, Coagulation of Salivary Mucoid by Freezing and Thawing of Saliva. Proceedings of the Society for Experimental Biology and Medicine 91, 565–569 (1956).

26. ThermoFisher Scientific, Pierce™ BCA Protein Assay Kit User Guide. (2161296), (2020).

27. C. L. Wardzala, A. M. Wood, D. M. Belnap, J. R. Kramer, Mucins Inhibit Coronavirus Infection in a Glycan-Dependent Manner. ACS Central Science 8, 351–360 (2022).

28. I. S. Woolhouse, D. L. Bayley, R. A. Stockley, Effect of sputum processing with dithiothreitol on the detection of inflammatory mediators in chronic bronchitis and bronchiectasis. Thorax 57, 667–671 (2002).

29. D. M. Kainz, B. J. Breiner, S. M. Früh, T. Hutzenlaub, R. Zengerle, N. Paust, Eliminating viscosity bias in lateral flow tests. Microsystems & Nanoengineering 7, 72 (2021).

30. G. A. Posthuma-Trumpie, J. Korf, A. van Amerongen, Lateral flow (immuno)assay: its strengths, weaknesses, opportunities and threats. A literature survey. Analytical and Bioanalytical Chemistry 393, 569–582 (2009).

31. W. Guo, J. Hansson, W. van der Wijngaart, Capillary Pumping Independent of Liquid Sample Viscosity. Langmuir 32, 12650–12655 (2016).

32. E. H. Yee, S. Lathwal, P. P. Shah, H. D. Sikes, Detection of Biomarkers of Periodontal Disease in Human Saliva Using Stabilized, Vertical Flow Immunoassays. ACS Sensors 2, 1589–1593 (2017).

33. I. B. Rodriguez-Calero, M. J. Coulentianos, S. R. Daly, J. Burridge, K. H. Sienko, Prototyping strategies for stakeholder engagement during front-end design: Design practitioners’ approaches in the medical device industry. Design Studies 71, 100977 (2020).

34. J. H. Shim, S. Y. Roh, J. S. Kim, D. C. Lee, S. H. Ki, J. W. Yang, M. K. Jeon, S. M. Lee, Normative measurements of grip and pinch strengths of 21st century korean population. Arch Plast Surg 40, 52–56 (2013).

35. N. H. L. Leung, Transmissibility and transmission of respiratory viruses. Nature Reviews Microbiology 19, 528–545 (2021).

36. R. Dhand, J. Li, Coughs and Sneezes: Their Role in Transmission of Respiratory Viral Infections, Including SARS-CoV-2. Am J Respir Crit Care Med 202, 651–659 (2020).

37. O. M. Allicock, M. E. Petrone, D. Yolda-Carr, M. Breban, H. Walsh, A. E. Watkins, J. E. Rothman, S. F. Farhadian, N. D. Grubaugh, A. L. Wyllie, Evaluation of saliva self-collection devices for SARS-CoV-2 diagnostics. medRxiv, (2021).

38. S. Y. Hwang, S. Y. Tan, P. T. Tan, C. Siau, Self-Swab and Saliva Collection for the Diagnosis of Covid-19. What Do Patients Feel About Them? Journal of Infectious Diseases and Epidemiology 6, (2020).

39. B. White, C. Day, H. H. Thein, A. Doab, A. Bates, J. Holden, I. Van Beek, L. Maher, Acceptability of hepatitis C virus testing methods among injecting drug users. Drug and Alcohol Review 27, 666–670 (2008).

40. D. A. Anderson, S. M. Crowe, M. Garcia, Point-of-Care Testing. Current HIV/AIDS Reports 8, 31–37 (2011).

41. S. R. Lee, G. D. Yearwood, G. B. Guillon, L. A. Kurtz, M. Fischl, T. Friel, C. A. Berne, K. W. Kardos, Evaluation of a rapid, point-of-care test device for the diagnosis of hepatitis C infection. Journal of Clinical Virology 48, 15–17 (2010).

42. G. Arora, S. Sheikh, S. Pallagatti, B. Singh, V. A. Singh, R. Singh, Saliva as a tool in the detection of hepatitis B surface antigen in patients. Compend Contin Educ Dent 33, 174–176, 178; quiz 180, 182 (2012).

43. H. Alqaderi, F. Hegazi, F. Al-Mulla, C.-J. Chiu, A. Kantarci, E. Al-Ozairi, M. Abu-Farha, S. Bin-Hasan, A. Alsumait, J. Abubaker, S. Devarajan, J. M. Goodson, H. Hasturk, M. Tavares, Salivary Biomarkers as Predictors of Obesity and Intermediate Hyperglycemia in Adolescents. Frontiers in Public Health 10, (2022).

44. Y. Z. Szabo, D. C. Slavish, Measuring salivary markers of inflammation in health research: A review of methodological considerations and best practices. Psychoneuroendocrinology 124, 105069 (2021).

45. R. K. Drobitch, C. K. Svensson, Therapeutic Drug Monitoring in Saliva. Clinical Pharmacokinetics 23, 365–379 (1992).

46. J. M. Yoshizawa, C. A. Schafer, J. J. Schafer, J. J. Farrell, B. J. Paster, D. T. Wong, Salivary biomarkers: toward future clinical and diagnostic utilities. Clin Microbiol Rev 26, 781–791 (2013).

47. Y. Gu, B. D. O. Shunmuganathan, X. Qian, R. Gupta, R. S. W. Tan, M. Kozma, K. Purushotorman, T. M. Murali, N. Y. J. Tan, P. R. Preiser, J. Lescar, H. Nasir, J. Somani, P. A. Tambyah, S. C. S. Group, K. G. C. Smith, L. Renia, L. F. P. Ng, D. C. Lye, B. E. Young, P. A. MacAry, Defining Factors that Influence vaccine-induced, cross-variant neutralizing antibodies for SARS-CoV-2 in Asians. medRxiv, 2022.2003.2006.22271809 (2022).

48. J. Park, An Optimized Colorimetric Readout Method for Lateral Flow Immunoassays. Sensors (Basel) 18, (2018).

