## Supplementary material for "SHEAR Saliva Collection Device Augments Sample Properties for Improved Analytical Performance": SHEAR SCD supplementary material

**This file includes:**

Figs. S1 to S4
Tables S1

### Supplementary figures

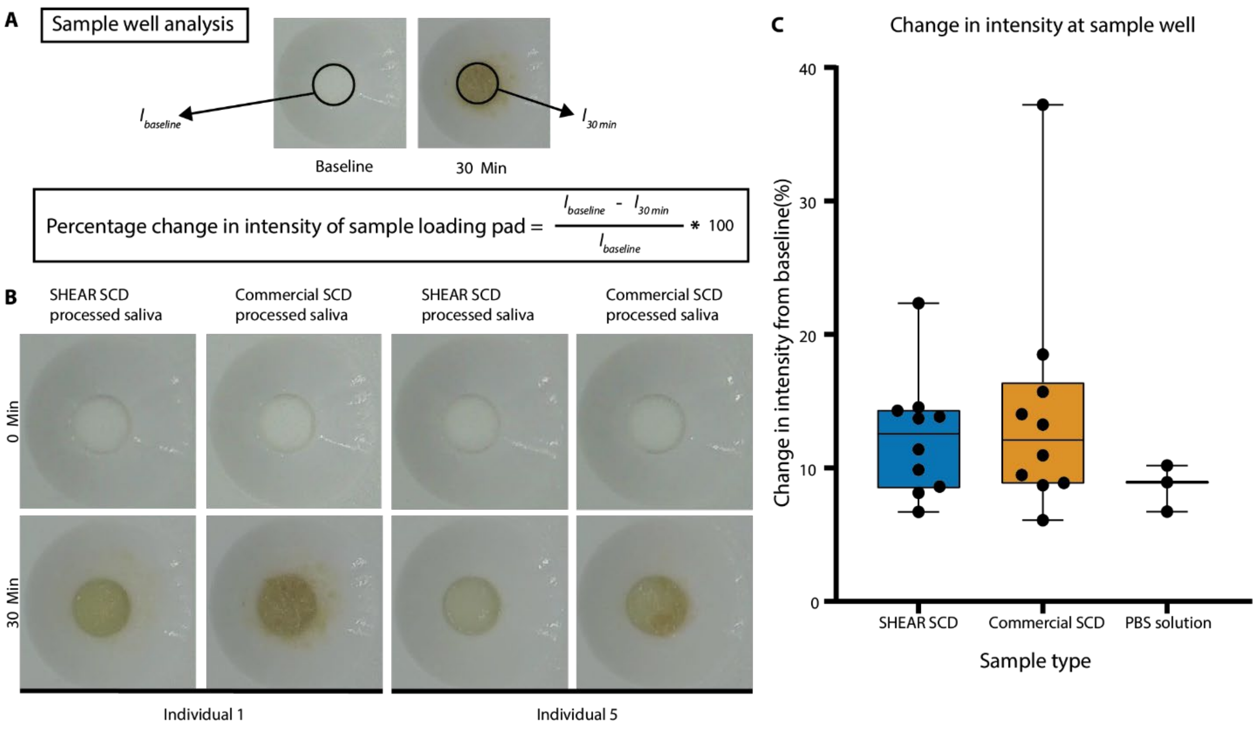


**FIG S1. Color intensity of sample well,**

(**A**). Analysis method of percentage change in colour intensity of the sample well. (**B**). Photograph of sample well of individual 1 and 5 at minute 0 and minute 30, before and after loading of SHEAR SCD processed and commercial SCD processed saliva samples respectively. (**C**). Change in colour intensity of the sample well of SHEAR SCD processed saliva (N=10), commercial SCD processed saliva (N=10) and PBS solution (N=3). Whiskers represent maximum and minimum values, and the box represents median value, 25th and 75th percentile. No statistical difference detected with Wilcoxon signed-rank test at $\alpha = 0.05$.

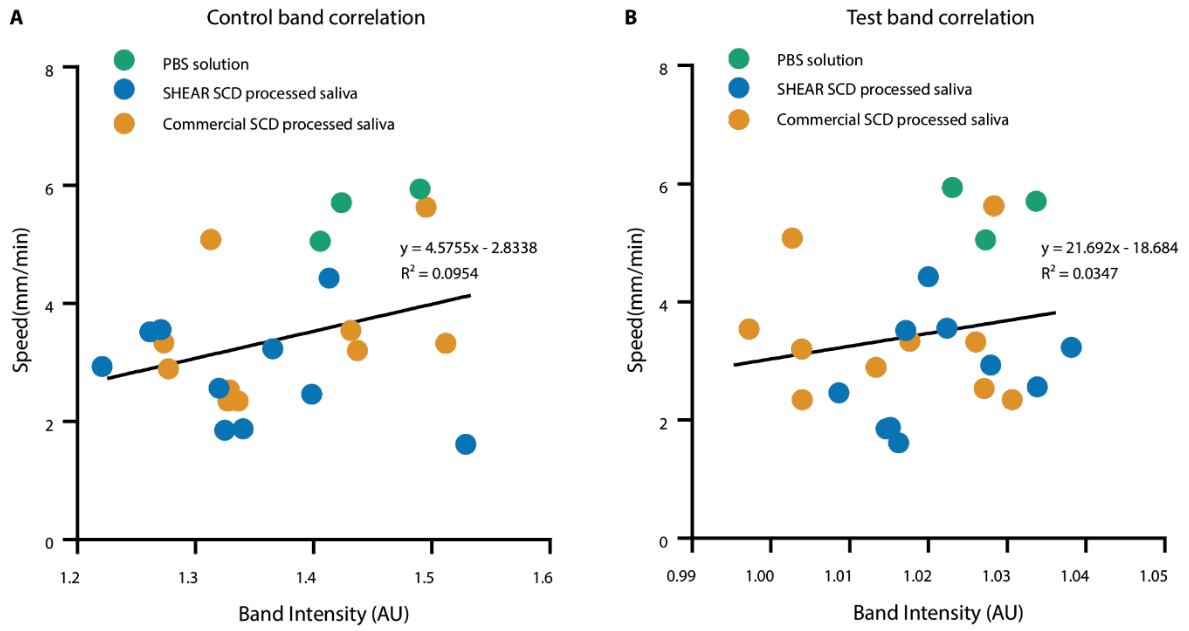


**FIG S2. Correlation between line intensity and liquid migration speed.**
Scatter plot with linear regression of liquid migration speed and intensity of (**A**) test line and (**B**) control line of SHEAR SCD processed saliva (N=10), commercial SCD processed saliva (N=10) and PBS solution (N=3).

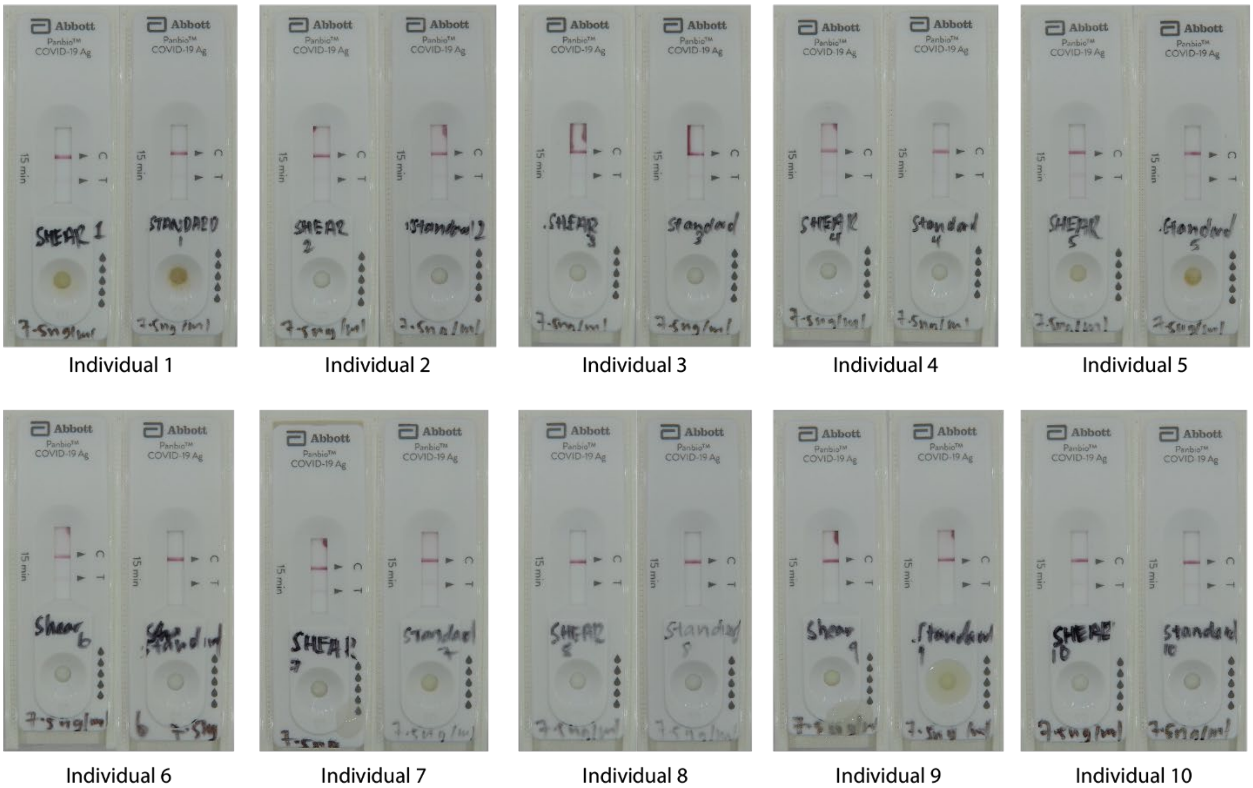


**FIG.S3. Photograph of rapid antigen test cassette used in the paired rapid antigen test.**

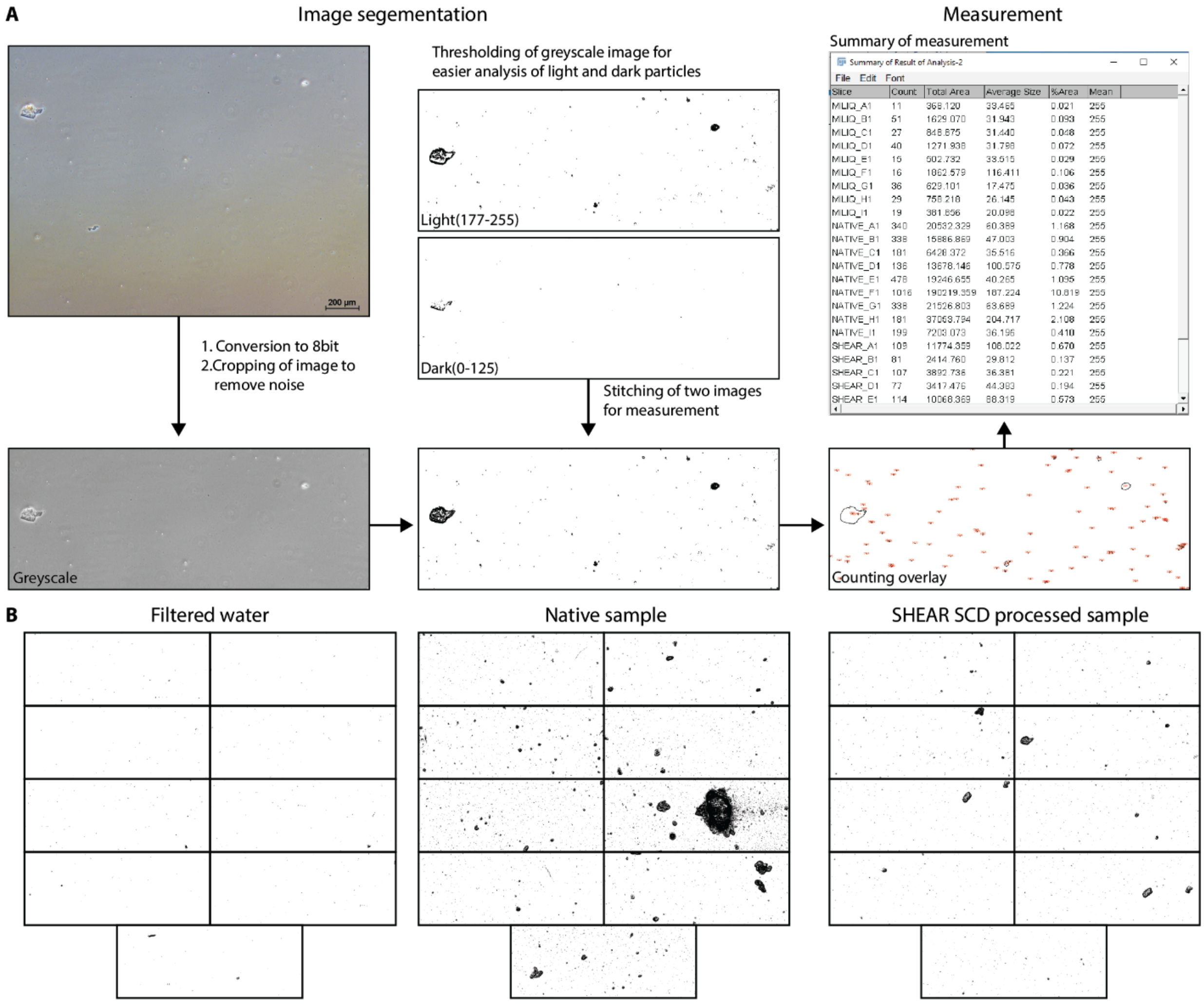


**FIG.S4. Processed image of for counting of food particle.**
(**A**). Method of image segmentation with ImageJ used for the measurement of area and count of food particles present in Filtered water, native sample and SHEAR SCD processed sample (N=9) (**B**). Processed images of Filtered water, native sample and SHEAR SCD processed sample

**Table S1. Example quotes of each focus area from the user study**

| **Focus area 1: Usability & Functionality of the SHEAR SCD** | | |
|---|---|---|
| | Specifics | Example quote |
| Positive | SHEAR device is straightforward and easy to use | "It was straightforward and it's easy to use to… in terms of how it functions, it's not hard to understand." – Participant 6 |
| | Wide funnel allows for easy spitting process | "Because there was a much larger surface area, [I] don't have to worry about trying to fit my mouth into the funnel in order to contribute the saliva." – Participant 5 |
| | Wide funnel feels safer due to reduction in saliva spillage | "I think [the SHEAR device] is safer because you [spit] closer. You put your mouth closer and you open the flaps. Nothing is missing and going outside." - Participant 2 |
| | Process of squeezing and rolling is quick and easy | "It was really easy, because I thought the saliva wouldn't go through but I just push it and it all went in one move. So, it was quite fast and efficient." – Participant 7 |
| | Process of squeezing and rolling allows for clearer indication of saliva level | "I thought that the rolling of the bag down to secrete the saliva down into the tube was quite interesting. I could see that after folding the bag and as I watch the saliva flow down, [I] could see the clear distinction between before being push and what's after; there are less bubbles." Participant 9 |
| | SHEAR device easier to use for elderly | "I find device 1 is better for those senior citizen because err, it can avoid spillage of the saliva" – Participant 12 |
| | Clear Instructions | "I think it is quite clear, once you go through the video, you are able to I mean know what to do with the device" Participant 14 |
| Negative | Commercial device more efficient and easier to use | "The [commercial] device is more straightforward because it is just like an opening and closing. You don't really need to squeeze it out but I was thinking if it is because the [SHEAR] device [has] a filter or something." – Participant 1 |
| | Difficulty with flap during the rolling process | "When I was doing the folding down part [the process] was a bit hard, so I just force it down. It didn't feel like there was a per… not perforation." – Participant 7 |
| | Difficulty fully opening funnel | "When you pull [the funnel] open, the top will open but the bottom doesn't open fully. So, the saliva gets stuck at somewhere in the middle where it open." – Participant 4 |
| | Uncertain about amount of saliva needed | "It wasn't very clear for me the extent to which [the saliva] was [dripping] down into the tube, so I didn't know if I needed to add more saliva into the upper portion the funnel." – Participant 6 |

| | | |
|---|---|---|
| | Unsealed funnel before rolling process | "I had trouble sealing the bag. I don't know if it's the… it just didn't seal, so I just folded it." – Participant 7 |
| | Backflow of saliva during the squeezing and rolling process | "The [SHEAR] device, there is a degree of backflow if you press too hard. I think there is too much air pressure inside the tube." Participant 10 |
| | People with disabilities might find SHEAR hard to use | "Some people particularly those who maybe are, have disabilities, it [SHEAR] might be quite hard to use" Participant 11 |
| **Focus area 2: Saliva as a biological material for diagnostic tests** | | |
| Positive | Increased comfort level | "It's [saliva collection] more comfortable than the nasal swab because that hurts" Participant 4 |
| | Saliva collection is simple to use | "Saliva [collection] will be the easiest and the most convenient method "Participant 10 |
| | Bleeding after swabbing | "But then the PCR swab was just uncomfortable and then I bleed afterwards, after the nurse poke my nose. It was not a good experience" Participant 10 |
| | Sneezing after nasal swab | "I would say that it is very uncomfortable to do the nasal swab because I will just keep sneezing after that" Participant 7 |
| | Prefers saliva collection over current collection methods | "I would definitely choose the saliva collection for sure, for the convenience, not too sure about the cost, but definitely how less invasive it is" Participant 3 |
| Negative | Safety concerns for saliva collection method | "But then the saliva is like you are putting… there is a risk of spillage, that's probably where my concern is." Participant 4 |
| | Prefer Cheek swab over saliva collection | "If I have a choice, I will rather do the cheek swab than do the saliva collection" Participant 7 |
| | Amount of saliva required for test | "I mean the only thing that I don't like about these procedures [Saliva collection] is you have to get a lot of saliva out of your throat" Participant 2 |
| **Focus area 3: User's consideration for the adoption of diagnostic kit with SHEAR SCD** | | |
| Device consideration | Accuracy | "If it is not more accurate, it wouldn't motivate me to use it. I would be like just give me any one [diagnostic kit] and I just use any one" Participant 7 |
| | Cost | "Price point relative to the ART test kits, as long as it doesn't stray too far, slightly expensive, I think it is worth the trade-off for being uncomfortable." Participant 9 |
| | Safety | "So, I guess mainly safety and … so that they are confident that it [diagnostic kit] will work and will be safe and will give a real result." Participant 2 |
| | Ease of use | "If I really have to choose between device [diagnostic kit], I care more about how easy it is to use, like how many steps" Participant 6 |
| | Hygiene | "I think the factors that would influence my… influence my preference to use this [diagnostic kit] would include things like hygiene" Participant 15 |

| | | |
|---|---|---|
| Saliva test requirements | Amount of saliva required | “Depends on the amount of saliva required. Saliva is preferred over swab test if no more than 1ml is required” Participant 11 |
| | Pre-testing requirements | “Then, I will probably have to figure out, like actually plan in advance when I want to take the [saliva] test because it also includes even if I take a small snack, … I can’t eat a small snack or brush my teeth, so when can I do it” Participant 5 |
